# Factors influencing survival outcomes in patients with stroke in Zimbabwe: A 12-month longitudinal study

**DOI:** 10.1101/2024.04.02.24305220

**Authors:** Farayi Kaseke, Lovemore Gwanzura, Cuthbert Musarurwa, Elizabeth Gori, Tawanda Nyengerai, Timothy Kaseke, Aimee Stewart

## Abstract

**Background:** In this longitudinal study, we aimed to determine factors influencing survival outcomes among patients with stroke over a 12-month period. The investigation sought to uncover influential determinants to enhance the precision of prognostic assessments and inform targeted interventions for individuals affected by strokes.

**Methods:** Employing a longitudinal study design, participants were observed for 12 months from baseline, censoring survivors at the endpoint. The dataset originated from a comprehensive study involving stroke patients treated at three referral hospitals in Zimbabwe: Parirenyatwa, Sally Mugabe, and Chitungwiza Central Hospital. The primary outcome variable, the duration of survival until death, was measured in days from the initiation of stroke treatment. Gompertz parametric regression analysis was utilized for data modeling following Accelerated Failure Time (AFT) model diagnostics.

**Results:** In our study, 188 stroke patients were enrolled at baseline. However, 51 patients were excluded from the analysis due to either missing information or loss to follow-up. Among the remaining 137 patients who were tracked over a 12-month period, 42% were censored, and 58% were deceased. Individuals utilizing ’Free Service (older than 65/pensioners/retirees)’ hospital bill payment methods showed a decreased risk of death (HR: 0.4, 95% CI: 0.20, 0.80), suggesting a protective effect compared to cash paying patients. Those with a secondary school level education displayed a significantly lower risk of death (HR: 0.2, 95% CI: 0.04, 0.69) compared to those without formal education. Age was a significant factor, with individuals aged 45-65 and those over 65 years showing higher adjusted hazard ratios (HR: 4.9, 95% CI: 1.80, 13.25; HR: 5.5, 95% CI: 1.92, 15.95, respectively) relative to those below 45 years of age. Housing status revealed a protective effect for those residing with parents/relatives (adjusted HR: 0.4, 95% CI: 0.20, 0.66), while individuals with a ’Very severe’ functional outcome showed an increased hazard (adjusted HR: 4.9, 95% CI: 1.12, 21.33).

**Conclusion:** The study findings demonstrate that hospital bill payment methods, housing status, educational attainment, functional outcome, and age significantly affect survival outcomes among stroke patients. This highlights the need to consider socio-demographic and clinical variables in the development of prognostic assessments and targeted interventions for individuals recovering from stroke.

## Introduction

Stroke, a significant global health challenge, is a major contributor to disability and mortality, particularly affecting low- and middle-income countries, where approximately 70% of stroke-related deaths and 87% of stroke-related disabilities occur (1). Within sub-Saharan Africa, the burden of stroke is disproportionately high, contributing significantly to stroke-related deaths and disability rates (2)(3). Alarming statistics reveal an annual incidence rate of up to 316 per 100,000, a prevalence of up to 1460 per 100,000, and a staggering 3-year mortality rate exceeding 80% (1).

Challenges in managing stroke in this region are further emphasized by Sahle Adeba et al. (2022), (4) who highlighted a high incidence of mortality among adult stroke patients. These complexities necessitate the need to understand factors influencing survival outcomes, thus providing a foundation for targeted interventions tailored to the unique socio-economic and healthcare landscape. Age consistently emerges as a pivotal factor influencing stroke outcomes, with individuals over 65 years facing a higher risk of death (5)(6). Furthermore, other studies highlighted the predictive role of old age in stroke incidence, emphasizing the importance of age-related considerations (7) (8). In addition, reduced educational attainment is associated with a heightened risk of mortality from brain stroke (9) (10). Unemployment, another socio-economic factor, has also been linked to increased mortality risk among stroke patients (11).

Recognizing the nature of stroke outcomes, it is important to delve into additional determinants such as hospital bill payment modalities, educational attainment, housing status, and total functional outcomes. This study saught to contribute to this understanding, by exploring factors influencing survival outcomes among stroke patients in Zimbabwe. By doing so, we aim to inform targeted interventions and enhance the overall management of stroke in this high-burden region, aligning with global efforts to reduce the impact of stroke on communities worldwide.

## Methods

### Study design

Our study employed a longitudinal design that initially enrolled 188 stroke patients at baseline. However, during the course of the 12-month follow-up, 51 patients were excluded from the analysis due to missing information or loss to follow-up. The exclusion of these 51 observations was meant to enhance the robustness and reliability of the analysis by focusing on participants with complete and reliable data. The subsequent analysis involved the remaining 137 patients who were followed-up for 12 months, from baseline, after which the surviving participants were censored. The dataset was derived from a larger study encompassing stroke patients initiating treatment at three referral hospitals (Parirenyatwa, Sally Mugabe, and Chitungwiza Central Hospital), in Harare, Zimbabwe. At all the three hospitals, the study participants were admitted in the stroke units or medical wards. All the hospitals were manned by neurologists, physiotherapists and occupational therapists who ran weekly outpatient stroke clinics for all stroke patients discharged from hospitals.

### Inclusion and exclusion criteria

All patients (age ≥ 18 years) diagnosed with stroke and admitted to the stroke units or the medical wards of the three public hospitals during the study period were included. The study excluded admitted patients who died before medical stabilization or refused to participate in the study as well as those had a Transient Ischaemic Attack (TIA).

### Study population

The source population was all stroke patients with a clinical diagnosis admitted to the three public referral hospitals in Harare, Zimbabwe from July 2015 to Nov 2017. Not all the patients had confirmatory diagnosis of stroke using a CT scan since some could not afford the cost. One hundred and eighty-eight adult stroke patients were included in the study.

### Study variables

The primary outcome variable was the duration of survival until death, measured in days from the initiation of stroke treatment to the occurrence of death or censorship. Various independent variables were examined for their potential influence on the survival time of stroke patients, including sex, hospital of admission, age group, marital status, occupational status, level of education, housing status, cigarette smoking status, hospital bill payment modality, transport modality, stroke side, comorbidities, time of stroke, total functional outcome, and dependency. This comprehensive set of variables allowed for a thorough exploration of factors contributing to the survival outcomes in the study population.

### Operational definitions

**Event**: death recorded among stroke patients during 12 months of follow-up (in the hospital or after hospital discharge in the period reported by a caregiver).

**Censored**: (i) if the patient was alive until the 365-day post stroke, (ii) if the patient died before they were medically stable (less than 72 hours post admission).

**Time to event**: Time from hospital admission until the death of the patient was confirmed either during admission or after discharge within the 12 months of follow-up.

**Twelve months case fatality rate**: was calculated using the number of deaths due to stroke during the 12- months follow-up period as the numerator and the total number of stroke patients during the 12 months follow-up period as the denominator

### Data collection procedure

Ethical clearance was obtained from The Parirenyatwa and University of Zimbabwe Joint Research Ethics Committee (JREC – 312/12) and The Medical Research Council of Zimbabwe (MRCZ – 34/78). All participants gave written informed consent and caregivers gave assent for those that could not communicate. Data collection was carried out by one research assistant for each hospital who was trained in the use of the data collection tools. The research assistants collected all relevant data from patients’ charts and interviewed the patients/caregivers using a validated data extraction sheet and questionnaire. Participant history was recorded from the patient and/or relatives using a language they were comfortable communicating in (Shona or English). Clinical information date was abstracted from the patients’ notes or files, Data on socio-demographics (age, sex, education level occupation, marital status etc), clinical data (clinical presentation, past medical illness, duration of symptoms, CT-scan etc) was collected by interview at baseline, while death was recorded upon follow up at 3 months or at 12 months. Finally, the participants were assessed for their overall functional outcomes using the Functional Independence Measure (FIM at baseline and at follow up. This was transformed into ordinal categories (mild, moderate, severe, and very severe) to generate proportions in each category. Fifty-one participants were lost to follow up or missing information, hence data for 137 participants were analyzed.

### Data analysis

The statistical analysis was conducted using STATA software version 15.1 (StataCorp, College Station, TX). Descriptive statistics were expressed as frequencies with corresponding percentages. The Kaplan Meier survival curves were utilized to compare patients by covariates. Bivariate Cox proportional hazards regression models were fitted for each explanatory variable. Variables with p≤0.25 in the bivariate Cox proportional hazards analysis were deemed eligible for further analysis. However, the Cox proportional hazards model failed the model diagnostics, as indicated by the global test result’s significance (p<0.05) and a systematic departure from a horizontal line in the plot of scaled Schoenfeld residuals against transformed time, revealing violations of proportional hazard assumptions. Consequently, Gompertz parametric regression analysis was employed for modeling the data post accelerated failure time (AFT) model diagnostics. Variables with a p-value < 0.05 were considered significantly associated with the outcome in the multivariate analysis, and the adjusted hazard ratios (HR) with a 95% confidence interval (CI) were utilized to express the associations between the outcome and independent variables. The Wald test was used to retain variables that had a p-value < 0.05, utilizing a process of iteratively removing, refitting, and verifying until all significant variables were retained in the final main effects model.

### Accelerated Failure Time Model Selection and Diagnostics

The AFT models were used since the Cox proportional hazards (PH) assumptions were violated. For an in-depth exploration of the impact of candidate covariates on the survival time of stroke patients, bivariate analysis was conducted for each covariate using different baseline distributions in AFT models. The multivariable analysis incorporated the Exponential, Gompertz, Weibull, and log-normal distributions for the baseline hazard function, considering the most significant covariates. To assess the goodness of fit, estimated cumulative hazard plots were plotted against Cox-Snell residuals, providing insight into how well the AFT models captured the stroke dataset. Furthermore, the Akaike’s information criterion (AIC) was employed to assess the model’s goodness of fit and guide the selection of the most appropriate AFT model for the analysis.

### Ethics approval and participant consent

Permission to carry out this study was granted by the institutional review boards at the three referral hospitals Parirenyatwa, Sally Mugabe (formerly Harare Central) and Chitungwiza Central hospitals. Ethical approval was given by the Joint Parirenyatwa Group of Hospitals and University of Zimbabwe Research Ethics Committee (JREC – 312/12) and The Medical Research Council of Zimbabwe (MRCZ – 34/78). All participants gave written informed consent and in the event that the participant could not communicate, assent was given by the caregiver.

## Results

### Sociodemographic characteristics

Table 1 presents the sociodemographic and clinical characteristics of stroke patients followed for 12 months. In total 79 (58%) participants were deceased and 58(42%) were censored. Sex distribution showed 83(61.3%) females and 54(38.7%) males, with corresponding deceased percentages of 57.1% and 58.5% for each sex. Hospital of admission showed variations, with Parirenyatwa having 33/47 (70.2%) of its patients deceased, Sally Mugabe Hospital with 22/54 (40.7%), and Chitungwiza Central Hospital with 24/36 (66.7%). Age group categorization revealed higher mortality in those over 65 years (80.8%), and marital status showed 58.2% deceased among married individuals. The ’Not in Union’ category, which included the single, widowed, divorced, and separated individuals, had 56.9% deceased individuals. The unemployed category had 61.3% deceased, while among retirees 73.1% of the participants died. Educational attainment also displayed variation in mortality with 100.0% deceased among those with no formal education and 78.3% deceased among those with only primary school level education.

**Table 1:** Sociodemographic characteristics of stroke patients followed for 12 months (n=137)

| Variable | Category | No. of patients | Survival Status |  |
| --- | --- | --- | --- | --- |
|  |  |  | Deceased (%) | Censored (%) |
| Sex | Female | 84 | 48(57.1) | 36(42.9) |
|  | Male | 53 | 31(58.5) | 22(41.5) |
| Hospital of Admission | Parirenyatwa Hospital | 47 | 33(70.2) | 14(29.8) |
|  | Sally Mugabe Hospital | 54 | 22(40.7) | 32(59.3) |
|  | Chitungwiza Central Hospital | 36 | 24(66.7) | 12(33.3) |
| Age Group (years) | < 45 | 28 | 7(25.0) | 21(75.0) |
|  | 45-65 | 57 | 30(52.6) | 27(47.4) |
|  | > 65 | 52 | 42(80.8) | 10(19.2) |
| Marital Status | Married | 79 | 46(58.2) | 33(41.8) |
|  | Not in Union** | 58 | 33(56.9) | 25(43.1) |
| Occupational Status | Unemployed | 75 | 46(61.3) | 29(38.7) |
|  | Employed / self-employed | 36 | 14(38.9) | 22(61.1) |
|  | Retired | 26 | 19(73.1) | 7(26.9) |
| Level of Education | No formal education | 3 | 3(100.0) | 0(0.0) |
|  | Primary | 60 | 47(78.3) | 13(21.7) |
|  | Secondary | 65 | 24(36.9) | 41(63.1) |
|  | Tertiary | 9 | 5(55.6) | 4(44.4) |
| Housing Status | Staying with parents / relatives | 36 | 17(47.2) | 19(52.8) |
|  | Own House | 80 | 51(63.8) | 29(36.2) |
|  | Tenant | 21 | 11(52.4) | 10(47.6) |
| Cigarette Smoking Status | Never smoked | 100 | 52(52.0) | 48(48.0) |
|  | Former | 16 | 13(81.2) | 3(18.8) |
|  | Current smoker | 21 | 14(66.7) | 7(33.3) |
| Hospital Bill Payment Modality | Free Service* | 19 | 11(57.9) | 8(42.1) |
|  | Cash | 104 | 60(57.7) | 44(42.3) |
|  | Health insurance | 11 | 6(54.6) | 5(45.4) |
|  | Social welfare | 3 | 2(66.7) | 1(33.3) |
| Transport Modality | Public | 117 | 66(56.4) | 51(43.6) |
|  | Private | 20 | 13(65.0) | 7(35.0) |
| Stroke Side | Bilateral | 1 | 0(0.0) | 1(100.0) |
|  | Left | 68 | 36(52.9) | 32(47.1) |
|  | Right | 68 | 43(63.2) | 25(36.8) |
| Comorbidities | No | 21 | 10(47.6) | 11(52.4) |
|  | Yes | 116 | 69(59.5) | 47(40.5) |
| Time of Stroke | Day | 93 | 53(57.0) | 40(43.0) |
|  | Night | 44 | 26(59.1) | 18(40.9) |
| Total Function | Mild | 6 | 2(33.3) | 4(66.7) |
|  | Moderate | 20 | 6(30.0) | 14(70.0) |
|  | Severe | 27 | 10(37.0) | 17(63.0) |
|  | Very severe | 84 | 61(72.6) | 23(27.4) |
| Dependency | Dependent | 131 | 77(58.8) | 54(41.2) |
|  | Independent | 6 | 2(33.3) | 4(66.7) |

Key: \*\* Not in Union includes, single, widowed, divorced and separated,

### Overall Kaplan-Meier estimation of survival

The overall Kaplan-Meier estimation of survival, as illustrated in Figure 1, indicates that the overall probability of survival was above 50% during the first 100 days but declined to below 50% by day 200. Over the 12-month follow-up period from baseline, involving a total of 137 patients with stroke, 42% of individuals survived, while 58% were deceased.

**Figure 1:**
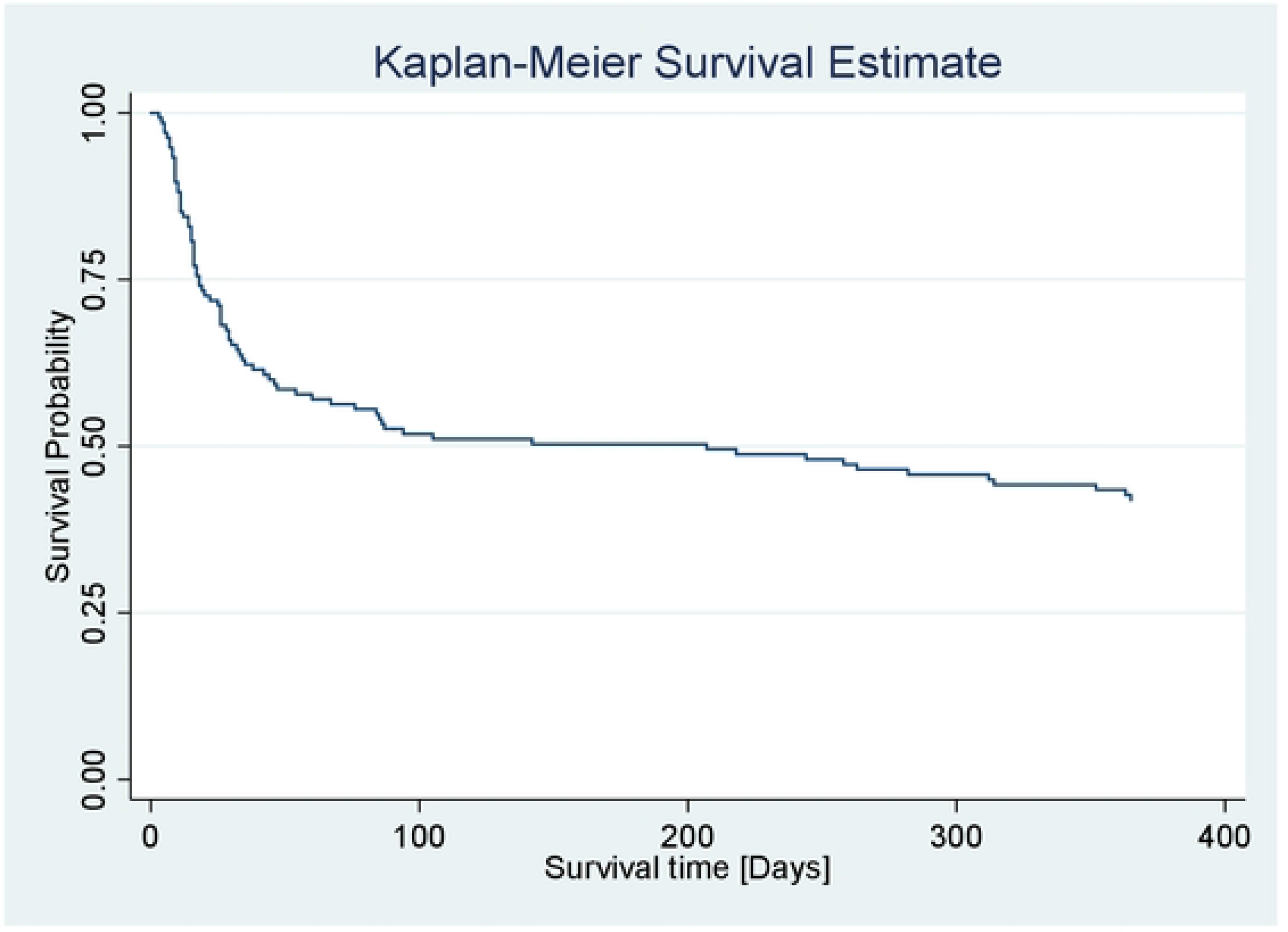
**Overall Kaplan-Meier survival status among stroke patients**

### Kaplan-Meier Survivor Estimates of Patients with stroke

Survival estimates for patients with stroke were analyzed using the Kaplan-Meier method to compare survival times. The results revealed distinct patterns based on functional outcomes. For instance, individuals with very severe functional outcomes at baseline had the shortest survival time, while those with mild and moderate outcomes survived the longest. Further comparison by age group demonstrated that patients below 45 years showed higher survival times, and the lowest survival times were observed in patients aged over 60 years. Additionally, based on participant housing status, patients staying with relatives or parents and those residing as tenants had longer survival times compared to those with own accommodation (Figure 2).

**Figure 2:**
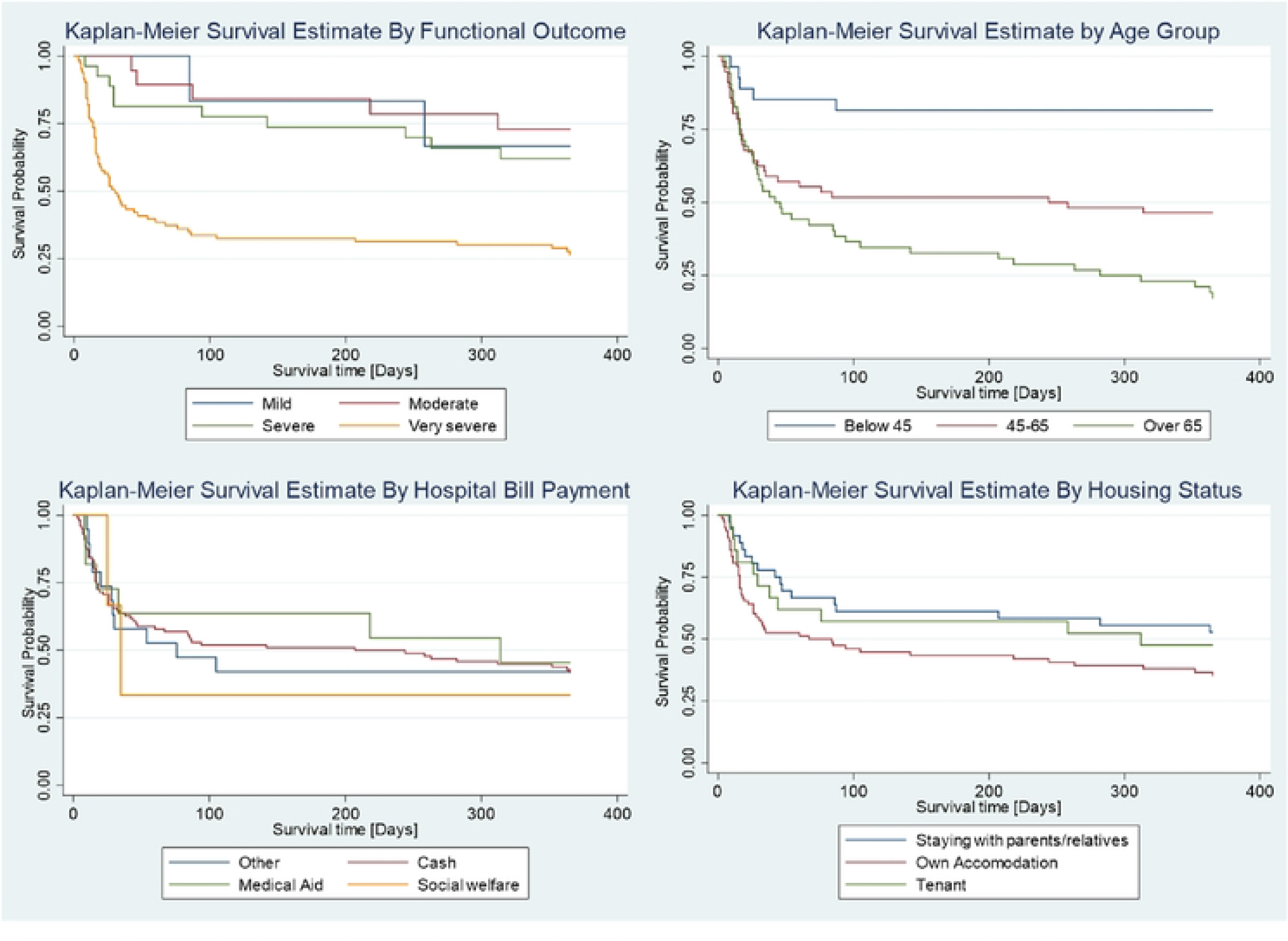
Kaplan-Meier survivor estimates by functional outcome, age group, hospital bill payment and housing status

### AFT Model Selection

The efficiency of the AFT models was compared using the Akaike Information Criterion (AIC). Among the alternative AFT models considered (Weibull, Exponential, Log-normal), the Gompertz AFT model showed the lowest AIC value (AIC = 400.3), signifying its best fit to the stroke dataset compared to other alternatives. Consequently, the Gompertz AFT model was chosen as the most suitable model for describing the characteristics of the stroke dataset (Table 2).

**Table 2:** Akaike’s information criterion values for parametric models.

| Model | AIC |
| --- | --- |
| Weibull | 414.4 |
| Exponential | 438.5 |
| Gompertz | 400.3 |
| Log-Normal | 403.5 |

Key: AIC-Akaike information criterion; BIC-Bayesian information criterion

### AFT Models Graphical Evaluation

The adequacy of model fit was further evaluated through graphical analysis using Cox-Snell residuals. Specifically, Cox-Snell residuals were employed to assess the overall goodness of fit across the different parametric models. Figure 3 illustrates the Cox-Snell residuals plots, revealing that the Gompertz AFT model demonstrated the most favorable fit for this dataset of stroke patients. This determination was based on the observation that the plot of Cox-Snell residuals against the cumulative hazard function of residuals formed an approximately straight line through the origin at a 45-degree angle, distinguishing it as the optimal fit when compared to the Exponential, Weibull, and log-normal models.

**Figure 3:**
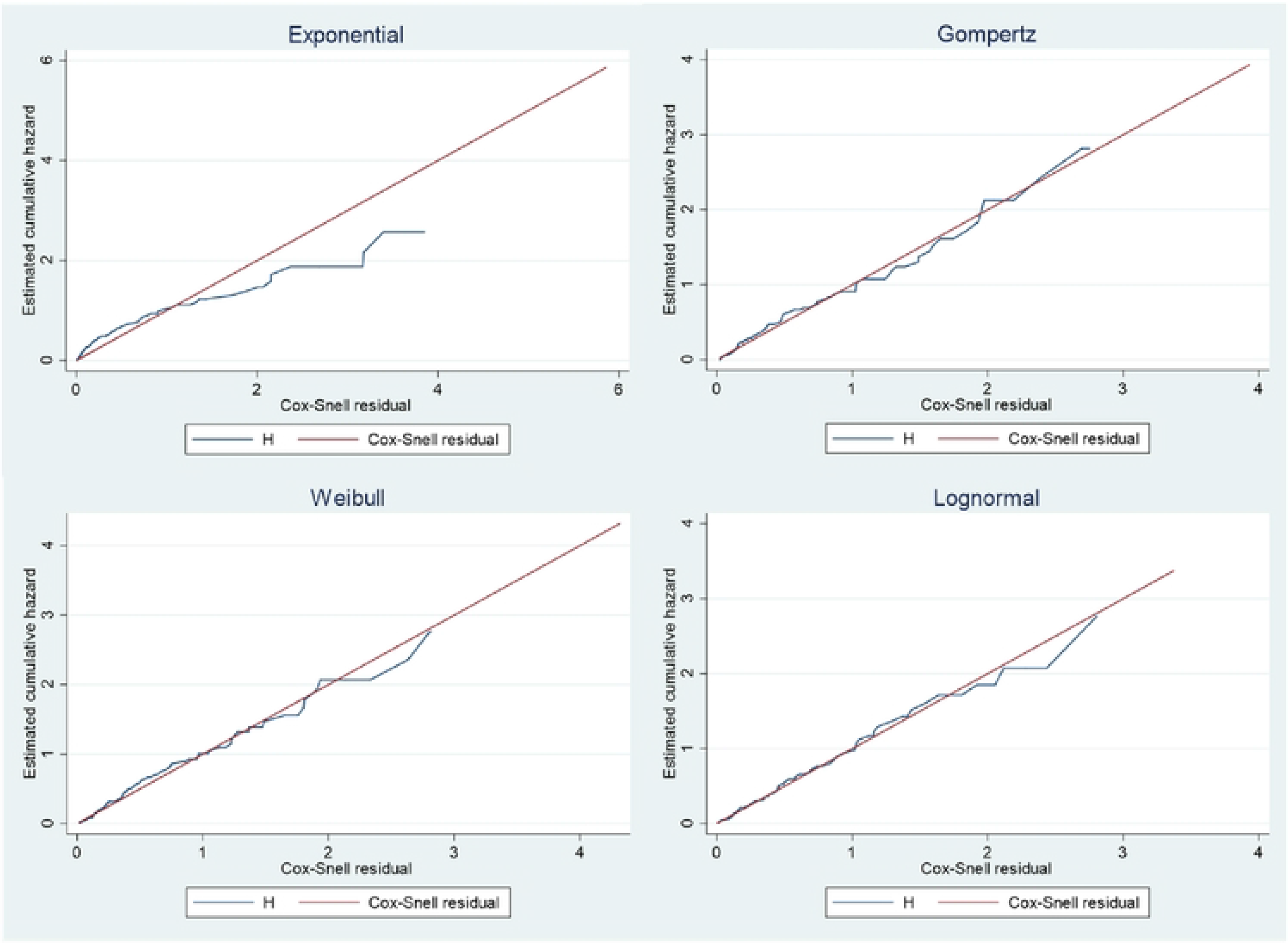
Estimated cumulative hazard against Cox-Snell residuals for the Exponential, Gompertz, Weibull and Lognormal models.

### Factors influencing survival among stroke patients

In the multivariable Gompertz AFT model for survival status among stroke patients (Table 3), several predictors were assessed. Regarding hospital bill payment modalities, compared to cash payments, those receiving free services which include older than 65, pensioners and retirees had a significantly lower risk of death 0.4(95%CI:0.20, 0.80), implying a protective effect on survival. However, the ’Health insurance’ and ‘Social Welfare’ modalities were not statistically significantly associated with survival. Educational attainment demonstrated a significant trend, with higher educational attainment associated with lower risk of mortality. For instance, individuals with secondary school level education had adjusted hazards ratios of 0.2(95% CI: 0.04, 0.69), indicating a significantly reduced risk compared to those with no formal education. Age was also significantly associated with mortality, with individuals aged 45-65years and those aged over 65 years showing adjusted hazards ratios of 4,9 (95%CI:1.80,13.25) and 5.5 (95%CI:1.92,15.95), respectively, compared to those below 45 years. Housing status showed a significantly protective effect for those staying with parents/relatives [0.4(95%CI: 0.20, 0.66)], and total functional outcome demonstrated a significantly increased hazard for individuals with a ’Very severe’ outcome [4.9 (95% CI: 1.12, 21.3)].

**Table 3:** Multivariable Gompertz AFT model for survival status and predictors among stroke patients (n=137).

| Variable | Category | Adjusted HR 95% CI |
| --- | --- | --- |
| Hospital Bill Payment modality | Cash | Reference |
|  | Free service | 0.4 (0.20, 0.80)** |
|  | Health insurance | 0.6 (0.26, 1.55) |
|  | Social welfare | 1.0 (0.24, 4.68) |
| Level of Education | No formal education | Reference |
|  | Primary | 0.4 (0.10, 1.31) |
|  | Secondary | 0.2 (0.04, 0.69)* |
|  | Tertiary | 0.2 (0.05, 1.21) |
| Age Group | < 45 | Reference |
|  | 45-65 | 4.9 (1.80, 13.25)** |
|  | > 65 | 5.5 (1.92, 15.95)** |
| Housing Status | Own House | Reference |
|  | Staying with parents / relatives | 0.4 (0.20, 0.66)*** |
|  | Tenant | 1.0 (0.48, 1.99) |
| Total Function | Mild | Reference |
|  | Moderate | 0.8 (0.15, 4.51) |
|  | Severe | 1.0 (0.20, 5.16) |
|  | Very severe | 4.9 (1.12, 21.3)* |

**Key:** \* p<0.05, \*\*p<0.01, \*\*\*p<0.001

## Discussion

This longitudinal study enrolled 188 stroke patients and sought to uncover influential determinants that influenced survival outcomes among stroke patients over a 12-month period. Overall, 79 (58%) stroke patients died during the 12 months’ period. This seemingly high proportion of deaths could be attributed to complications that develop among stroke patients over time as well as complications of bed rest. This finding is however, in concordance with global trends as between 20 to 75% per cent of first-time patients with stroke reported die within a month, and a third by 6 months (12; 13). Overall case fatality rates have been reported rage from 18% to 62% in some African countries (13;14). Contrasting results showing much lower death rates of 2.5% in the first year of follow-up were reported in Thailand (12). The reasons for this low mortality rate are not explained.

Although varied, the mortality rates in different African countries could be due to limited health care facilities and uncontrolled risk factors, poor functional abilities post-stroke, older age and impaired consciousness (14–17). Another explanation could be that most patients could not afford to go to the hospital collect medications due to a lack of finances before stroke. Thus once they suffered a stroke, their conditions got worse being furthers compounded by inability to travel or fund medications (18, 19).

Findings from the present study, provide valuable insights into the factors influencing survival outcomes among stroke patients. We previously reported an in-hospital fatality rate of 25% in a one-year retrospective study at these three same hospitals (20). The multivariable Gompertz Accelerated Failure Time model allowed for a comprehensive assessment of various predictors, shedding light on the influence of socio- demographic and clinical variables on survival time of individuals’ post-stroke.

Educational attainment emerged as a consistent and significant predictor of mortality post stroke, unveiling a compelling trend where higher educational attainment correlated with a reduced risk of death among stroke patients. Particularly noteworthy was the substantially lower risk observed in individuals with secondary school level education compared to those with no formal education, and this is similar to previous research findings (9) (10). This finding highlights the pivotal role of education as a social determinant of health, influencing factors such as health literacy, access to resources, and possibly adherence to medical recommendations. As such, efforts to enhance educational opportunities and health literacy may contribute to improved survival outcomes among stroke patients.

Age was an influential determinant. Individuals aged 45-65 and over 65years showed substantially higher hazard ratios compared to those below 45 years. This also aligns with existing studies (5)(6), emphasizing the impact of age on stroke outcomes and reinforces the important need for tailored care strategies that consider the unique needs and challenges associated with different age groups. Tailoring interventions based on age-specific considerations may enhance the effectiveness of stroke care and improve overall survival rates.

Housing status emerged as another determinant, revealing a protective effect for those staying with parents/relatives. This suggests a potential role for social support and familial environments in mitigating mortality risk among stroke patients. Understanding the dynamics of familial support systems and their impact on patient outcomes could inform the development of interventions that leverage social support networks to enhance overall stroke care.

Total functional outcome, a key aspect of stroke management, surfaced as a significant predictor in this study. Individuals classified as having ’Very severe’ stroke showed a substantially increased hazard of death. This demonstrates the critical importance of assessing and addressing functional outcomes in stroke management, as severe functional impairment significantly amplifies the risk of mortality. Tailoring interventions to address functional limitations and promote rehabilitation may contribute to improved survival outcomes for stroke patients. Previous studies have also highlighted an association between functional impairment, specifically cognitive impairment, and an elevated risk of mortality among individuals affected by stroke (21).

Hospital bill payment modalities played a significant role in influencing survival outcomes. Notably, individuals receiving free services, which include pensioners, retirees, and those older than 65, showed a considerably lower risk of death compared to those making cash payments. Patients in this category receive hospital services without charge, as long as the services are available at public hospitals. In contrast, individuals relying on cash payments may face challenges obtaining services due to financial constraints, potentially contributing to higher death rates.

This suggests a potentially protective effect associated with alternative payment methods. The lack of significant associations with Health Insurance’ and ‘Social Welfare’ payment modalities prompts further exploration in these payment systems and their differential impact on survival outcomes. These modes of payment may depend on the government’s ability to sustain them; otherwise, individuals under these categories may also be treated similarly as cash paying patients. In addition, some of the required services may not be available within public hospitals, and this would compel patients to seek such services outside the public health system.

### Strengths and limitations of the study and or findings

The strengths of the study were: recruitment was from central hospitals improved generalisability, Prospective cohort design, a fairly large baseline sample size, among others. The findings of the present study highlighted the factors associated with mortality in this group of patients and paves the way for health professionals to develop guidelines to mitigate mortality among stroke patients. The limitations are attributed to missing information about the cause of death and inability to get the information thereby introducing potential selection bias despite efforts to minimize it. This led to analysis only for the available data with a substantially diminished sample size thus statistical analysis on causality could not be carried out. Our study was only on patients who were admitted in the hospitals and did not include those who died before admission and those who were not admitted due to various reasons. Wide confidence intervals on some variables suggest a possible need for replication of the study with much larger sample size. The 12- month follow-up might have limited capturing long-term survival patterns, potentially missing late-stage events and ongoing health challenges beyond this timeframe.

### Conclusions

The observed influence of hospital bill payment modalities on survival outcomes, with receiving free hospital services associated with a lower risk of death, suggesting potential avenues for healthcare policy and practice improvements. Additionally, the consistent and significant trend in educational level underscores its role as a crucial social determinant of health in the context of stroke, influencing health literacy, resource access, and adherence to medical recommendations. The critical impact of age on survival outcomes emphasizes the need for tailored care strategies for different age groups. Furthermore, the protective effect of staying with parents/relatives and the heightened mortality risk associated with ’Very severe’ functional outcomes emphasize the importance of assessing and addressing social support systems and functional impairment in stroke management. These findings highlight the nature of stroke prognosis, emphasizing the need for patient-centered approach in stroke care that considers and addresses diverse determinants impacting survival outcomes. Community based studies may give more information about case fatality rates among stroke patients in Zimbabwe.

## Data Availability

Data is available upon request.

## Acknowledgements

Firstly, we would like to acknowledge the participants of the study. Our special thanks and appreciation also goes to the research assistants and staff of the three public referral hospitals; Parirenyatwa, Sally Mugabe and Chitungwiza Central Hospitals in Zimbabwe. We would also want to acknowledge the funders of the study - NIH Fogarty student support grant (Grant Number D43TW009539).

